# Deep Neural Survival Networks for Cardiovascular Risk Prediction: The Multi-Ethnic Study of Atherosclerosis (MESA)

**DOI:** 10.1101/2021.04.12.21255286

**Authors:** Quincy A. Hathaway, Naveena Yanamala, Matthew J. Budoff, Partho P. Sengupta, Irfan Zeb

## Abstract

**Background:** There is growing interest in utilizing machine learning techniques for routine atherosclerotic cardiovascular disease (ASCVD) risk prediction. We investigated whether novel deep learning survival models can augment ASCVD risk prediction over existing statistical and machine learning approaches.

**Methods:** 6,814 participants from the Multi-Ethnic Study of Atherosclerosis (MESA) were followed over 16 years to assess incidence of all-cause mortality (mortality) or a composite of major adverse events (MAE). Features were evaluated within the categories of traditional risk factors, inflammatory biomarkers, and imaging markers. Data was split into an internal training/testing (four centers) and external validation (two centers). Both machine learning (COXPH, RSF, and lSVM) and deep learning (nMTLR and DeepSurv) models were evaluated.

**Results:** In comparison to the COXPH model, DeepSurv significantly improved ASCVD risk prediction for MAE (AUC: 0.82 vs. 0.79, *P*≤0.001) and mortality (AUC: 0.86 vs. 0.80, *P*≤0.001) with traditional risk factors alone. Implementing non-categorical NRI, we noted a 65% increase in correct reclassification compared to the COXPH model for both MAE and mortality (*P*≤0.05). Assessing the relative risk of participants, DeepSurv was the only learning algorithm to develop a significantly improved risk score criteria, which outcompeted COXPH for both MAE (4.07 vs. 2.66, *P*≤0.001) and mortality (6.28 vs. 4.67, *P*=0.014). The addition of inflammatory or imaging biomarkers to traditional risk factors showed minimal/no significant improvement in model prediction.

**Conclusion:** DeepSurv can leverage simple office-based clinical features alone to accurately predict ASCVD risk and cardiovascular outcomes, without the need for additional features, such as inflammatory and imaging biomarkers.

## 1.1 Introduction

Coronary artery disease is the leading cause of morbidity and mortality in the United States [1]. There are multiple algorithms available such as the Framingham risk score (FRS) [2], Reynold risk score [3, 4], and Pooled Cohort Equation (PCE) [5] for risk stratification and to help guide preventive strategies. These algorithms use traditional risk factors to derive an individual’s cardiac risk. Traditional risk factors have commonly either underestimated or overestimated an individual’s risk of coronary disease, thus promoting a search for subclinical markers. Detection of subclinical calcified atherosclerosis, such as coronary artery calcium (CAC), has been shown to improve the performance of these risk prediction models [6-8]. While subclinical markers have provided additive benefit to risk prediction, there remains a limited understanding regarding the incremental value of these biomarkers to allow for cost-effective clinical predictions [9].

Traditionally, survival models have been used to investigate the relationships between patients’ features and adverse outcomes, such as cardiovascular events, mortality, etc. [10-12]. Standard survival models like the Cox proportional hazards model (COXPH) employ several linear assumptions that allow it to predict outcomes for an individual patient. While COXPH utilizes a standard statistical approach, it can be evaluated in the context of training and testing sets to provide predictive values and function as a machine learning algorithm. Recently, a modern COXPH model implementing deep neural networks, referred to as DeepSurv, has been suggested as an alternative method for solving survival analysis problems [13]. DeepSurv has the capacity to create more complex and abstract feature combinations by utilizing hidden layers. These hidden layers provide independent nodes of neural network development, allowing the algorithm to exhaust the potential of features and feature combinations for event prediction. Additionally, DeepSurv is equipped with gradient descent optimization, providing an efficient means of minimizing the error of the algorithm through continual optimization of its parameters [13].

In this study we illustrate the value of DeepSurv over traditional models, such as COXPH, for predicting adverse cardiovascular events in the multiethnic study of atherosclerosis (MESA). Further, we assess the additive value of feature combinations by integrating traditional risk factors, inflammatory biomarkers, and imaging markers for deriving optimal risk stratification algorithms. Ultimately, we suggest a framework for the development, and use, of deep neural network risk scores in clinical medicine.

## 1.2 Methods

### 1.2.1 Implementation of The Multi-Ethnic Study of Atherosclerosis (MESA)

The MESA is a prospective study examining 6,814 men and women in the United States, with participants across an age range of 45 – 84 years old. The study was designed to include a multiethnic population of White/Caucasian (38%), Black/African American (28%), Hispanic (23%), and Chinese American (11%) participants. In the current study, information from participants obtained during the first exam (between the years 2000-2002) were used as features for event prediction using machine learning and deep learning approaches **(Figure 1A)**. The primary outcomes included myocardial infarction, resuscitated cardiac arrest, congestive heart failure, coronary revascularization, or all-cause mortality as major adverse events (MAE). Time of the event was classified as death (mortality) or the time at which the first cardiovascular outcome variable was detected (MAE). Only participants without clinical cardiovascular disease at baseline were included in the study.

**Figure 1:**
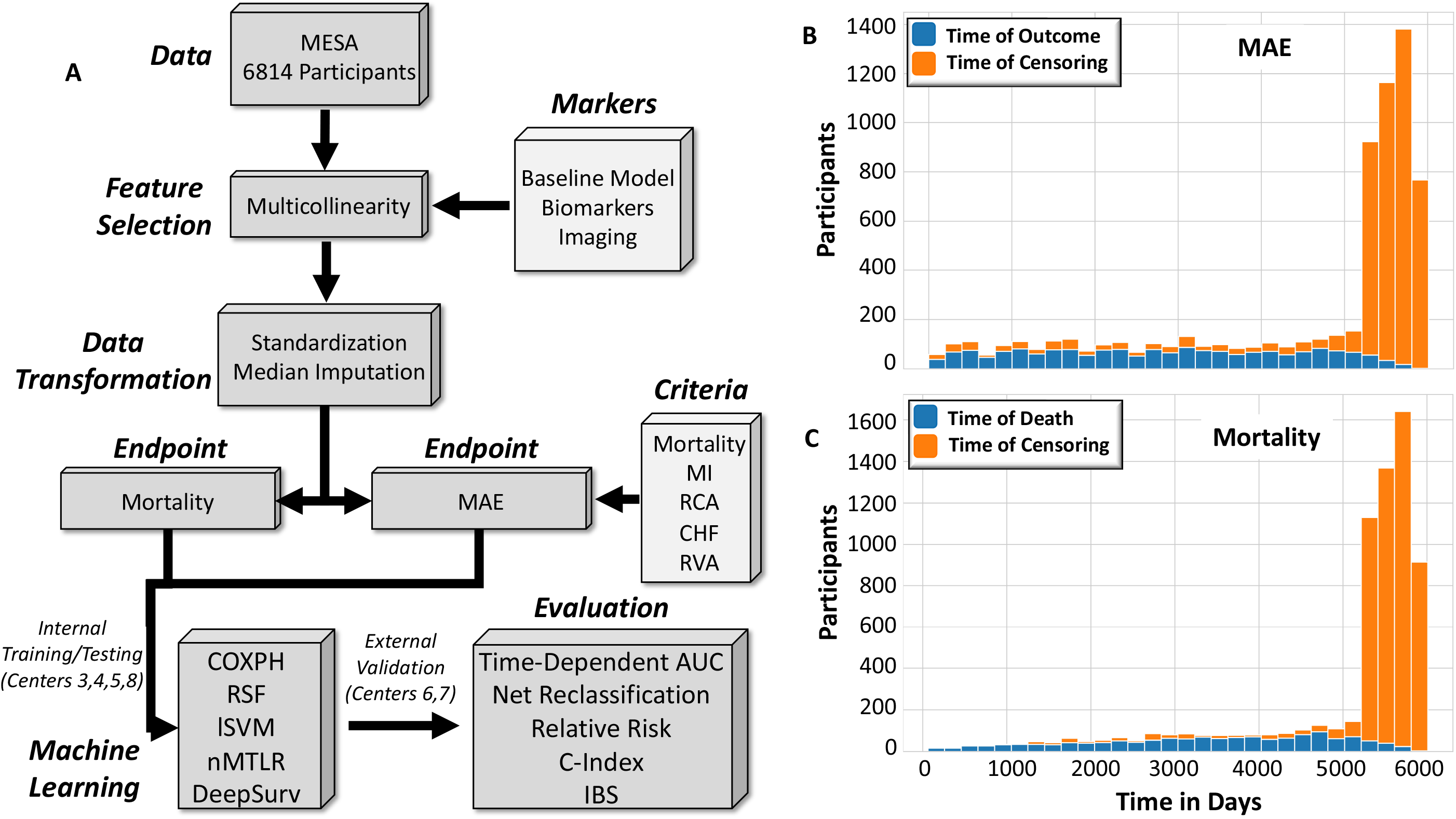
Methodological and data considerations for the Multi-Ethnic Study of Atherosclerosis (MESA). **(A)** Schematic detailing the flow of information through the survival analysis pipeline. Briefly, features from 6,814 participants in the MESA cohort were selected from three major categories: traditional risk factors (Baseline Model), inflammatory biomarkers (Biomarkers), and left ventricular structure/function (Imaging). Features exhibiting multicollinearity were removed, data was standardized and imputed. Classification of endpoints (mortality and MAE) were trained (internal training/testing set) and tested (external validation) using the machine learning models COXPH, RSF, and lSVM and deep learning models nMTLR and DeepSurv. Outcomes were evaluated through the cumulative/dynamic AUCs (Time-Dependent AUC), net reclassification improvement (NRI), relative risk (RR), concordance index (CI), concordance index with inverse probability of censoring weights (IPCW), and integrated brier score (IBS). **(B)** Data distribution along the survival timeline. Censored participants for mortality (78.7%) and MAE (72.3%) are displayed in orange and those with outcome/death in blue. MAE = major adverse events, including all-cause mortality and cardiovascular outcomes, Time in Days = time in days to the predicted event, COXPH = Cox Proportional Hazards Regression Model, RSF = Random Survival Forest, lSVM = Linear Support Vector Machines, nMTLR = Neural Multi-Task Logistic Regression, DeepSurv = Non-Linear Cox Proportional Hazards Deep Neural Network.

### 1.2.2 Feature Selection

To assess the strength of event prediction in the presence of features obtained from different clinical modalities, we selected features from three major categories, including traditional risk factors (Baseline Model), inflammatory biomarkers (Biomarkers), and left ventricular morphology, pericardial fat, and coronary artery calcium (CAC) (Imaging). Features included in each category were selected based on the following: 1) the feature was measured/assessed in greater than 70% of the participant cohort, 2) the feature has been shown to be correlated with clinical or subclinical findings, and 3) features exhibiting multicollinearity (correlation coefficient > 0.80) were removed except for one in each set of correlations **(Supplemental Figure 1)**. The R package corrplot 0.84 was used for multicollinearity correlations [14].

We utilized a linear proportional hazards model, without machine learning, to examine concordance index on the entire dataset. Imaging features provided higher concordance index scores (0.77 vs. 0.75 Baseline Model) for MAE, whereas Biomarkers provided higher concordance index scores (0.79 vs. 0.77 Baseline Model) for predicting mortality **(Supplemental Figure 2A)**. The feature combinations suggest that factors shaping the ultrastructure of the left ventricle and coronary atherosclerosis, are the most influential toward correctly identifying MAE, while inflammatory biomarkers markers (homocysteine, IL6, plasmin antiplasmin, etc.) are involved more prominently with understanding mortality, which supports previous findings [7, 10, 15-17]. The R package pec 2019.11.03 provided concordance indexes, and curves, through COXPH [18].

### 1.2.3 Data Transformation and Data Distribution

All non-binary features were given a normal, standard distribution using the preprocessing function in scikit-learn 0.23.1 [19]. Missing values were imputed with the median value of each column through the mlr 2.17.1 package in R 3.6.2 [20]. Data was split into an internal training/testing (∼66%, 4,584 participants) and external validation (∼33%, 2,230 participants) set. The internal training/testing and external validation split was determined by the field center in which the participants were enrolled. In order to derive an external validation set of ∼33%, field centers six (St. Paul, Minnesota) and seven (Chicago, Illinois) were allocated to the external validation group while field centers three (Forsyth County, North Carolina), four (New York, New York), five (Baltimore City and Baltimore County, Maryland), and eight (Los Angeles County, California) were selected as the internal training/testing set. Demographic information for the internal training/testing and external validation sets are provided **(Table 1)**.

**Table 1:** Internal Training/Testing and External Validation Set Demographics. Demographics and feature distribution of internal training/testing and external validation datasets. Data are statistically significant with a *P* ≤ 0.05 when denoted by a “**\***”. Data are represented as the mean and the 95% confidence interval. MAE = major adverse events, including all-cause mortality and cardiovascular outcomes.

| Features | Internal Training/Testing (Centers 3,4,5,8) N = 4,584 [95% CI] | External Validation (Centers 6,7) N = 2,230 [95% CI] | P-Value |
| --- | --- | --- | --- |
| **Traditional Risk Factors** |  |  |  |
| Age (Years) | 62.52 [62.23-62.82] | **61.39 [60.96-61.82]\*** | **≤0.001** |
| Gender (Male, %) | 46.71 [45.26-48.15] | 48.07 [46.00-50.15] | 0.289 |
| BMI (kg/m<sup>2</sup>) | 28.49 [28.33-28.65] | **28.02 [27.80-28.25]\*** | **≤0.001** |
| Education Status (0-8 scale) | 4.52 [4.46-4.59] | **5.20 [5.10-5.30]\*** | **≤0.001** |
| Income Category (1-13 scale) | 8.15 [8.05-8.26] | **9.15 [9.00-9.30]\*** | **≤0.001** |
| Race/Ethnicity (%) |  |  |  |
| 1. White/Caucasian | 31.85 [30.50-33.20] | **52.11 [50.03-54.18]\*** | **≤0.001** |
| 2. Chinese American | 10.89 [9.98-11.79] | **13.68 [12.25-15.10]\*** | **≤0.001** |
| 3. Black, African American | 34.69 [33.31-36.06] | **13.54 [12.12-14.96]\*** | **≤0.001** |
| 4. Hispanic | 22.58 [21.37-23.79] | 20.67 [18.99-22.35] | 0.075 |
| Diabetic (%) | 14.77 [13.74-15.80] | **10.54 [9.26-11.81]\*** | **≤0.001** |
| Kidney Dysfunction (%) | 10.12 [9.25-11.00] | **8.16 [7.02-9.30]\*** | **0.010** |
| Hypertensive (%) | 47.08 [45.63-48.52] | **37.00 [34.99-39.00]\*** | **≤0.001** |
| Hyperlipidemic (%) | 38.53 [37.12-39.93] | **36.01 [34.02-38.00]\*** | **0.044** |
| Metabolic Syndrome (%) | 37.46 [36.05-38.86] | **33.14 [31.18-35.10]\*** | **≤0.001** |
| Smoking Status (%) |  |  |  |
| 1. Never | 51.64 [50.19-53.08] | **48.12 [46.04-50.19]\*** | **0.006** |
| 2. Former | 35.49 [34.11-36.88] | **38.57 [36.54-40.59]\*** | **0.014** |
| 3. Current | 12.87 [11.90-13.84] | 13.32 [11.91-14.73] | 0.607 |
| Family History of Heart Disease (%) | 42.61 [41.13-44.09] | 43.04 [40.93-45.15] | 0.744 |
| **Inflammatory Biomarkers** |  |  |  |
| Homocysteine (μmol/L) | 9.33 [9.23-9.44] | 9.33 [9.16-9.50] | 0.586 |
| IL6 (pg/mL) | 1.58 [1.54-1.62] | 1.52 [1.47-1.57] | 0.054 |
| Plasmin Antiplasmin (nM) | 4.82 [4.76-4.88] | 4.75 [4.65-4.85] | 0.237 |
| Fibrinogen Antigen (mg/dL) | 348.1 [346.0-350.3] | **344.1 [341.1-347.1]\*** | **0.021** |
| C Reactive Protein (mg/L) | 3.88 [3.71-4.05] | **3.58 [3.33-3.82]\*** | **≤0.001** |
| D Dimer (μg/mL) | 0.38 [0.36-0.41] | 0.36 [0.33-0.40] | 0.447 |
| Factor VIII (%) | 100.8 [99.65-101.9] | **95.78 [94.31-97.24]\*** | **≤0.001** |
| **Left Ventricular Structure and Function** |  |  |  |
| LV Ejection Fraction < 50 (%) | 1.37 [1.04-1.71] | 0.99 [0.58-1.40] | 0.176 |
| Left Ventricular Area (mm<sup>2</sup>) | 4034 [4013-4055] | **3985 [3955-4015]\*** | **0.013** |
| Pericardial Fat (mm<sup>2</sup>) | 78.18 [76.99-79.36] | **82.47 [80.64-84.31]\*** | **≤0.001** |
| Total Agatston Calcium Score (Phantom Adjusted) | 145.7 [133.4-158.0] | 148.1 [131.1-165.1] | 0.924 |
| **Outcome Variables** |  |  |  |
| MAE (%) | 28.40 [27.10-29.71] | 26.23 [24.41-28.06] | 0.060 |
| Mortality (%) | 22.19 [20.98-23.39] | **18.70 [17.08-20.33]\*** | **≤0.001** |

### 1.2.4 Machine and Deep Learning Models and Evaluation

The framework for implementing and evaluating the machine and deep learning models is provided in the *Supplemental Materials and Methods*. Event prediction and feature importance were evaluated in three machine learning models (Cox Proportional Hazards Regression Model (COXPH), Random Survival Forest (RSF), Linear Support Vector Machines (lSVM)) and two deep learning models (Neural Multi-Task Logistic Regression (nMTLR) and Non-Linear Cox Proportional Hazards Deep Neural Network (DeepSurv)). Computations were performed in Python 3.7, all code is provided: https://github.com/qahathaway/MESA.

### 1.2.5 Statistics

GraphPad Prism 8.4.3 and RStudio 3.6.2 were used for statistical analyses. For assessing two continuous variables, a two-tailed, unpaired Student’s t-test was employed when normally distributed. Normality was tested using the Shapiro-Wilk test. If non-Gaussian distribution, two continuous variables were assessed using the Mann Whitney test. For analyses of more than one group of continuous variables, a one-way analysis of variance (ANOVA) was used. Confidence intervals were calculated for ROC curves through the combined Wilson/Brown method. Relative risk (RR) was calculated through the Koopman asymptotic score. Significance of RR, positive predictive value (PPV), negative predictive value (NPV), and AUCs were determined through log transformation of mean and confidence intervals using a z-score distribution [21]. All data was considered statistically significant if the *P*-value was less than or equal to 0.05 (*P* ≤ 0.05). Data are reported as the mean with the 95% confidence interval (CI).

## 1.3 Results

### 1.3.1 Characteristics of the Internal Training/Testing and External Validation Sets

The data was split into an internal training/testing (∼66%) and external validation (∼33%) set, which was determined by the field center/county in which the participants were assessed **(Table 1)**. Both the external validation and internal training/testing sets were of similar age, though statistically different (61.39 vs. 62.52 years old, *P*≤0.001), with no differences in gender (48.07% vs. 46.71% male, *P*=0.289). The external validation set compared to the internal training/testing set contained a higher percentage of White/Caucasians (52.11% vs. 31.85%, *P*≤0.001) and Chinese Americans (13.68% vs. 10.89%, *P*≤0.001), with a lower percentage of Black/African Americans (13.54% vs. 34.69%, *P*≤0.001). The external validation set exhibited a lower risk of experiencing mortality (18.70% vs. 22.19%, *P*≤0.001) but similar risk of MAE (26.23% vs. 28.40%, *P*=0.060) compared to the internal training/testing set, respectively. Of note, coronary artery calcium (CAC) was not significantly different in the external validation group (148.1 Phantom Adjusted) compared to the internal training/testing (145.7 Phantom Adjusted) (*P*=0.924).

### 1.3.2 Evaluation Metrics of Model Performance and Prediction Accuracy

The cumulative/dynamic AUC provides a means for plotting the ROC curve at any time throughout the survival analysis to assess predictions on which participants have, and do not have, the outcome at that time. With traditional risk factors, only linear Support Vector Machines (lSVM) and the Non-Linear Cox Proportional Hazards Deep Neural Network (DeepSurv) had significantly higher AUCs for both MAE and mortality in the Baseline Model and for All Features, compared to COXPH **(Table 2)**. DeepSurv had the highest performance criteria across all feature combinations and endpoints. Compared to COXPH, DeepSurv provided significant improvement with traditional risk factors for MAE (0.82 vs. 0.79, *P*≤0.001) and mortality (0.86 vs. 0.80, *P*≤0.001) as well as improved prediction with all features for MAE (0.85 vs. 0.81, *P*≤0.001) and mortality (0.87 vs. 0.84, *P*≤0.001). DeepSurv further outperforms COXPH in defining outcomes in racial/ethnic groups, indicated by prediction of MAE for White/Caucasians (0.82 vs. 0.80, *P*=0.005), Chinese Americans (0.82 vs. 0.77, *P*=0.001), Black/African Americans (0.77 vs. 0.74, *P*≤0.001), and Hispanics (0.81 vs. 0.78, *P*≤0.001) and mortality for White/Caucasians (0.85 vs. 0.83, *P*=0.034), Chinese Americans (0.84 vs. 0.70, *P*≤0.001), Black/African Americans (0.81 vs. 0.75, *P*≤0.001), and Hispanics (0.88 vs. 0.81, *P*≤0.001), using traditional risk factors alone **(Supplemental Table 1)**.

**Table 2:**
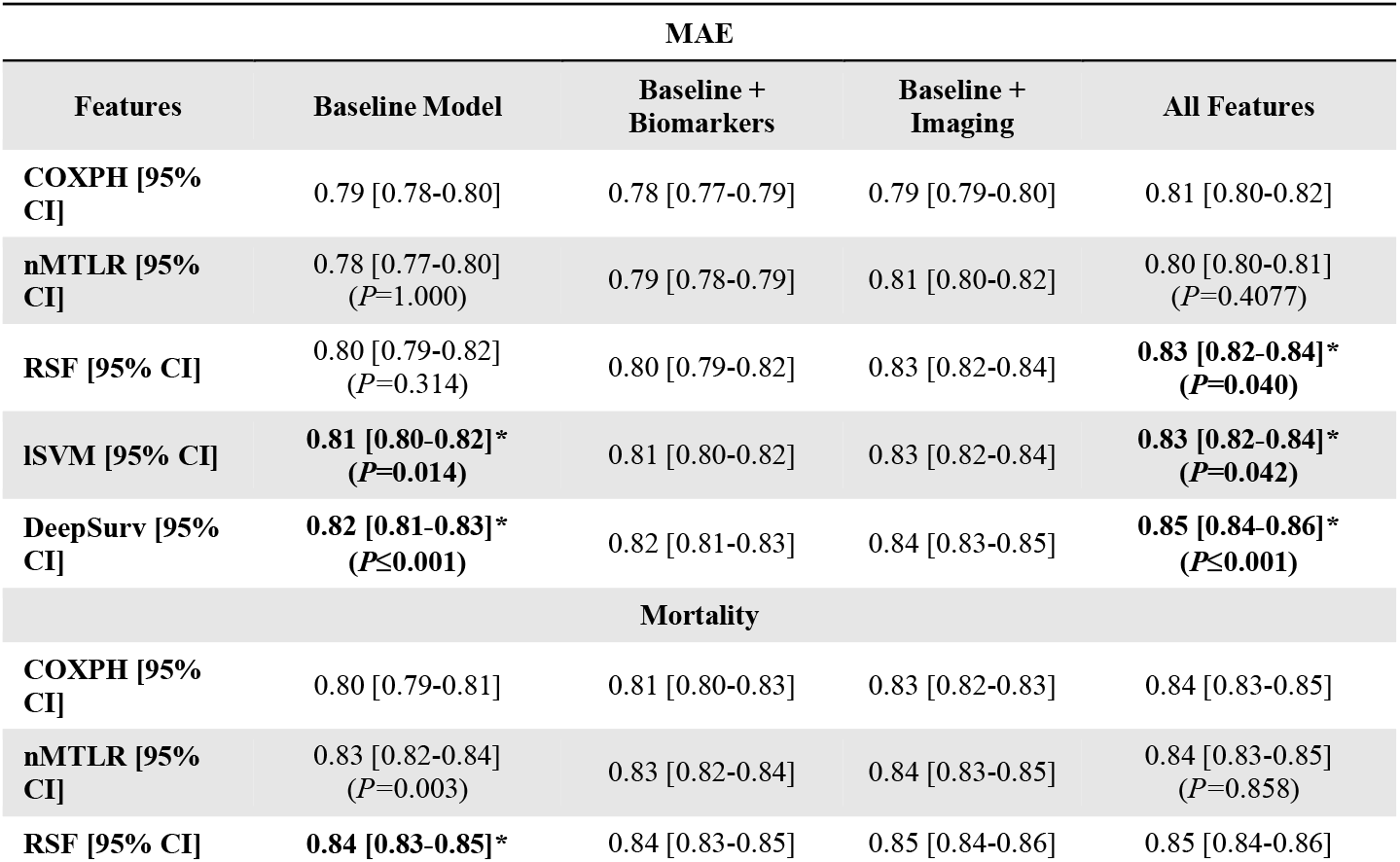
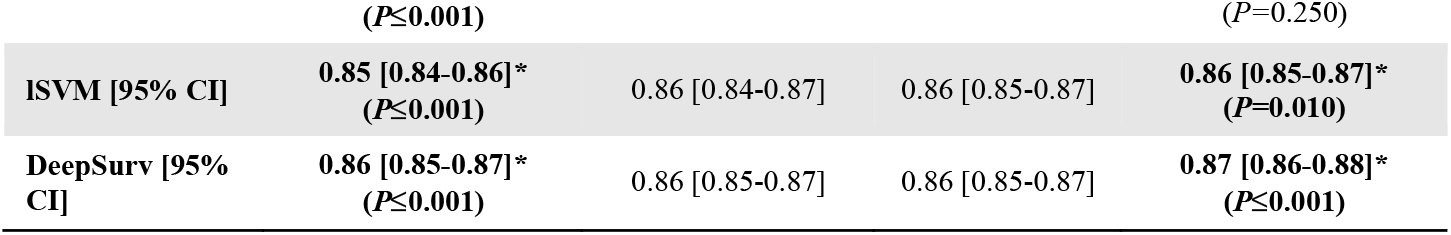
Cumulative/Dynamic Time-Dependent AUC. Cumulative/dynamic time-dependent AUCs for the COXPH, RSF, lSVM, nMTLR, and DeepSurv models. Data are statistically significant with a *P* ≤ 0.05 when denoted by a “**\***”. Data are represented as the mean and the 95% confidence interval. MAE = major adverse events, including all-cause mortality and cardiovascular outcomes, Baseline Model = traditional risk factors, Biomarkers = inflammatory biomarkers, Imaging = left ventricular structure and function, All Features = traditional risk factors, inflammatory biomarkers, and left ventricular structure and function, COXPH = Cox Proportional Hazards Regression Model, RSF = Random Survival Forest, lSVM = Linear Support Vector Machines, nMTLR = Neural Multi-Task Logistic Regression, DeepSurv = Non-Linear Cox Proportional Hazards Deep Neural Network.

The use of concordant pairs in survival analyses provides a metric to determine the point along the survival curve at which a subject is predicted to experience an outcome. Examining the concordance index for traditional risk factors, only DeepSurv performed significantly better in predicting concordant pairs **(Supplemental Table 2)** and concordant pairs adjusted for the inverse probability of censoring weights (IPCW) **(Supplemental Table 3)** for both MAE and mortality. DeepSurv provided the highest concordance score for MAE (0.78) and mortality (0.83) for traditional risk factors compared to COXPH (0.74 and 0.78, respectively). The integrated brier score **(Supplemental Table 4)** indicates the differences between predicted and observed probabilities, which was generally similar between machine and deep learning models for MAE and mortality.

### 1.3.3 Feature Importance

To determine which features contributed to the composition of each of the machine and deep learning models, permutation importance (COXPH, lSVM, nMTLR, DeepSurv) and mean decrease Gini (RSF) were employed **(Figure 2)**. When examining all of the features, participant age was one of the top five predictors for predicting MAE and mortality. CAC was one of the top five predictors in all models predicting MAE, while biomarkers of inflammation were more important for predicting mortality **(Figure 2)**. To further understand how individual features contributed to the construction of the machine and deep learning models, the best and worst predictive features were assessed.

**Figure 2:**
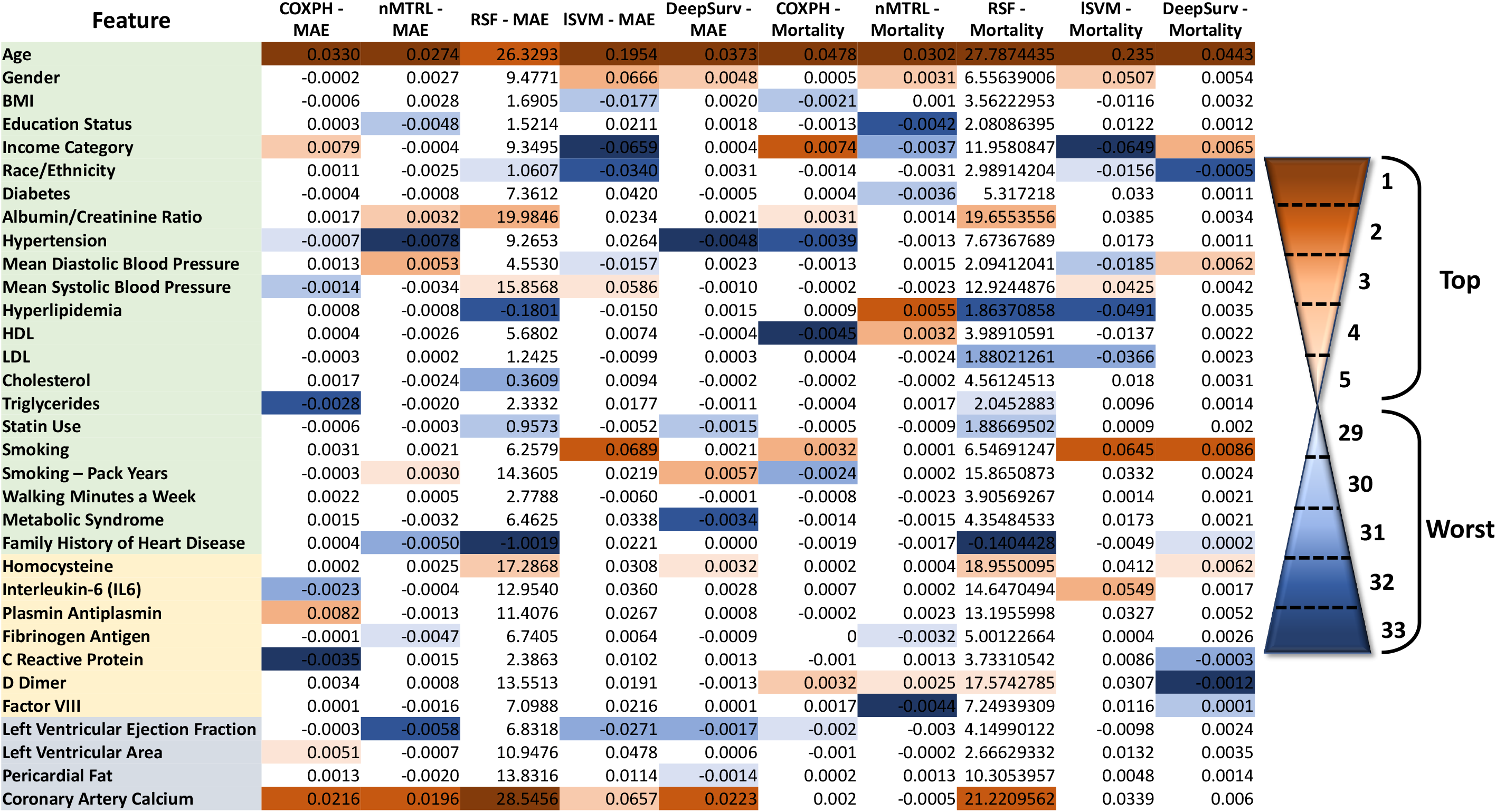
Feature importance for the COXPH, RSF, lSVM, nMTLR, and DeepSurv models. Permutation importance was applied to COXPH, lSVM, nMTLR, and DeepSurv, while mean decrease Gini (RSF) was implemented for RSF. The top five (orange) and worst five (blue) predictive features are shown for each model. MAE = major adverse events, including all-cause mortality and cardiovascular outcomes, COXPH = Cox Proportional Hazards Regression Model, RSF = Random Survival Forest, lSVM = Linear Support Vector Machines, nMTLR = Neural Multi-Task Logistic Regression, DeepSurv = Non-Linear Cox Proportional Hazards Deep Neural Network.

The top five performing features across all models with non-binary values were selected **(Figure 2)** with each model being trained on the internal training/testing set and fit with the external validation set. The cumulative/dynamic AUC for MAE **(Supplemental Figure 4)** and mortality **(Supplemental Figure 5)** for the top five performing features revealed minimal changes across all models. This suggests that the utility of these features are being completely exhausted by each of the machine and deep learning algorithms, with little, to no, room for improvement. To understand if features which comprise the worst predictors for each model performed similarly to the top predictors, the cumulative/dynamic AUCs were repeated for MAE **(Supplemental Figure 6)** and mortality **(Supplemental Figure 7)**. In this case, there were marked differences between the ROC curves for mortality and MAE, with the AUCs of COXPH (0.46 and 0.44), RSF (0.54 and 0.55), lSVM (0.55 and 0.55), nMTLR (0.55 and 0.55), and DeepSurv (0.56 and 0.56) revealing diverse handling of the predictors, respectively.

### 1.3.4 Predicted Risks for Clinical Decision Making

Machine and deep learning models were used in event prediction and risk stratification of patients by generating a risk score from the predicted risks of each model. Non-categorical net reclassification improvement (NRI) applied to the risk scores of traditional risk factors highlights the improved reclassification of nMTLR, RSF, lSVM, and DeepSurv compared to COXPH for MAE and mortality **(Table 3)**. DeepSurv had the highest percent reclassification and correctly classified cases for MAE (65% and 67%, *P*≤0.05) and mortality (65% and 72%, *P*≤0.05), While lSVM had the highest percent correctly classified controls (69% and 69%, *P*≤0.05) for MAE and mortality, respectively. Similar reclassification was seen when applied to all features **(Supplemental Table 5)**. To determine why DeepSurv can better identify patients who succumbed to MAE or mortality, we performed a categorical NRI using cutoffs derived from the Pooled Cohort Equation (PCE) estimated 10-year risk for atherosclerotic cardiovascular disease (ASCVD); this allowed for defining estimated risk score cutoffs of 5% (0.27, low risk), 7.5% (0.36, intermediate risk), and 20% (0.78, high risk). DeepSurv provided the most significant reclassification between the low risk and intermediate risk categories for MAE and mortality **(Supplemental Table 6)**, suggesting that DeepSurv can better identify individuals with outcomes in this “borderline” category than can COXPH.

**Table 3:**
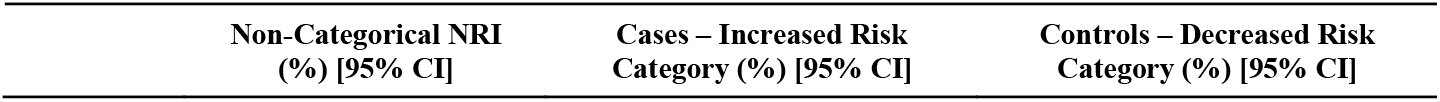
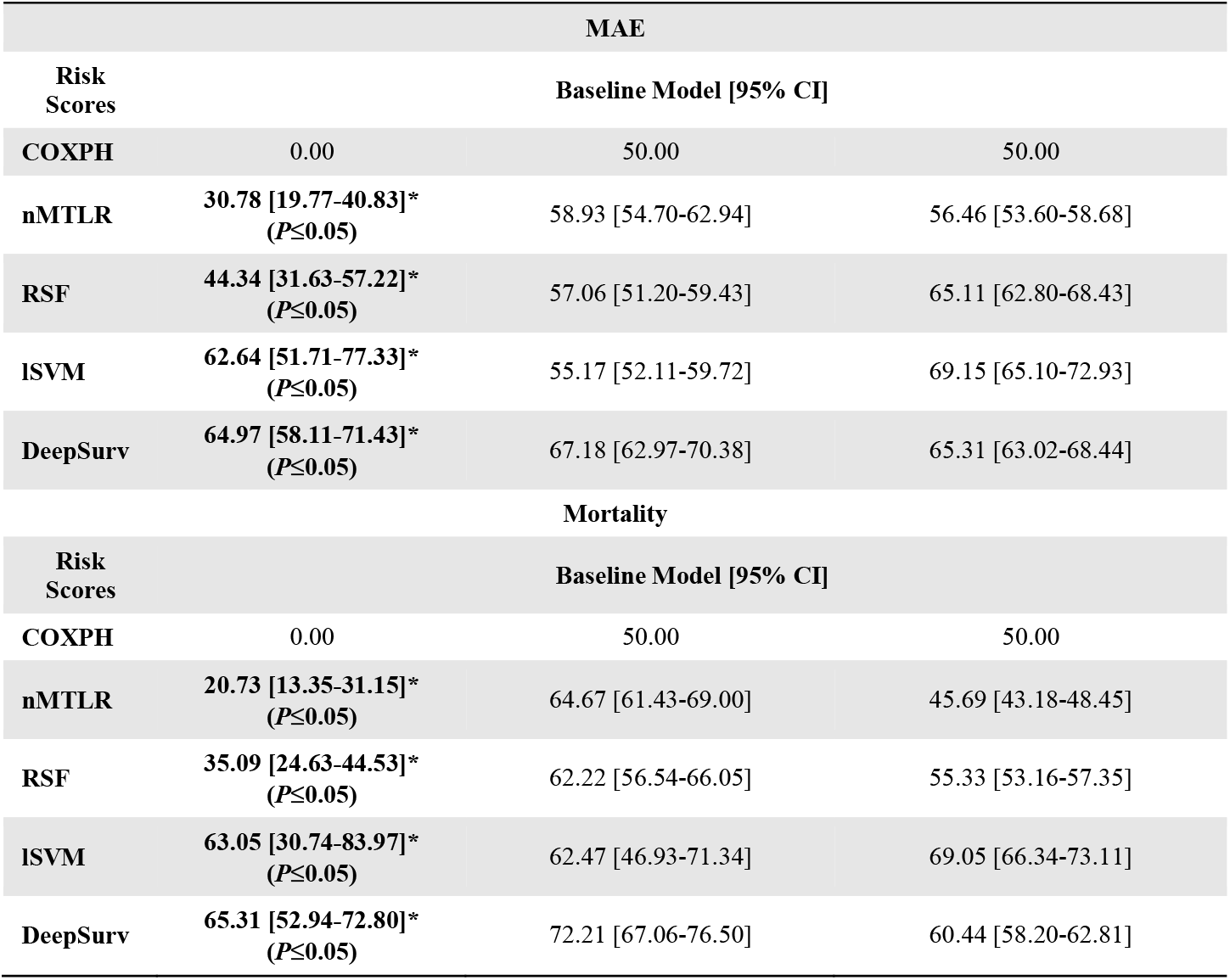
Reclassification – Baseline Model. Non-categorical net reclassification improvement (NRI) and the percentage of participants moving up and down risk categories for the COXPH, RSF, lSVM, nMTLR, and DeepSurv models. Analyses performed on the Baseline Model. Data are statistically significant with a *P* ≤ 0.05 when denoted by a “**\***”. Data are represented as the mean and the 95% confidence interval. MAE = major adverse events, including all-cause mortality and cardiovascular outcomes, Cases = participant with an event, Controls = participants without an event, Baseline Model = traditional risk factors, COXPH = Cox Proportional Hazards Regression Model, RSF = Random Survival Forest, lSVM = Linear Support Vector Machines, nMTLR = Neural Multi-Task Logistic Regression, DeepSurv = Non-Linear Cox Proportional Hazards Deep Neural Network.

The PCE ASCVD risk score classifies individuals with a score greater than 20% as being at high-risk for an event in the subsequent 10 years [22]. Using this criterion, 20% was set as a cutoff for determining relative risk (RR) of patients for mortality and MAE events in the machine and deep learning model risk scores. Using traditional risk factors alone, DeepSurv provided the highest increasing in RR and positive predictive value (PPV) for MAE (4.07 vs. 2.66, *P*≤0.001, and 66% vs. 51%, *P*≤0.001) and mortality (6.28 vs. 4.67, *P=*0.014, and 57% vs. 50%, *P=*0.027), compared to COXPH **(Table 4)**. Similar RR, PPV, and negative predictive value (NPV) were seen when applied to all features **(Supplemental Table 7)**.

**Table 4:** Predicted Risk – Baseline Model. Relative risk (RR), positive predictive value (PPV), and negative predictive value (NPV) for the COXPH, RSF, lSVM, nMTLR, and DeepSurv models. Analyses performed on the Baseline Model. Data are statistically significant with a *P* ≤ 0.05 when denoted by a “**\***”. Data are represented as the mean and the 95% confidence interval. MAE = major adverse events, including all-cause mortality and cardiovascular outcomes, Baseline Model = traditional risk factors, COXPH = Cox Proportional Hazards Regression Model, RSF = Random Survival Forest, lSVM = Linear Support Vector Machines, nMTLR = Neural Multi-Task Logistic Regression, DeepSurv = Non-Linear Cox Proportional Hazards Deep Neural Network.

|  | Relative Risk [95% CI] | Positive Predictive Value (%) [95% CI] | Negative Predictive Value (%) [95% CI] |
| --- | --- | --- | --- |
| **MAE** |  |  |  |
| **Risk Scores** | **Baseline Model [95% CI]** |  |  |
| **COXPH** | 2.66 [2.33-3.02] | 51.48 [46.98-55.95] | 80.63 [78.71-82.41] |
| **nMTLR** | 3.28 [2.89-3.72]* ($P=0.025$) | 58.32 [53.77-62.72]* ($P=0.036$) | 82.22 [80.37-83.83] ($P=0.220$) |
| **RSF** | 3.57 [3.14-4.02]* ($P \leq 0.001$) | 60.86 [56.35-65.19]* ($P=0.004$) | 82.94 [81.11-84.62] ($P=0.076$) |
| **ISVM** | 3.91 [3.42-4.47]* ($P \leq 0.001$) | 58.67 [54.59-62.64]* ($P=0.021$) | 84.98 [83.18-86.62]* ($P \leq 0.001$) |
| **DeepSurv** | 4.07 [3.59-4.61]* ($P \leq 0.001$) | 65.71 [61.22-69.94]* ($P \leq 0.001$) | 83.85 [82.07-85.49]* ($P=0.013$) |
| **Mortality** |  |  |  |
| **Risk Scores** | **Baseline Model [95% CI]** |  |  |
| **COXPH** | 4.67 [3.96-5.49] | 49.67 [45.09-54.26] | 89.36 [87.84-90.71] |
| **nMTLR** | 5.02 [4.26-5.92]<br>(*P*=0.541) | 50.87 [46.32-55.40] (*P*=0.716) | 89.87 [88.38-91.19] (*P*=0.619) |
| **RSF** | 5.90 [4.98-6.97]<br>(*P*=0.051) | 54.59 [50.01-59.09] (*P*=0.137) | 90.74 [89.30-92.00] (*P*=0.170) |
| **ISVM** | 5.87 [4.85-7.10]<br>(*P*=0.074) | 48.38 [44.35-52.42] (*P*=1.000) | **92.03 [90.62-93.24]\* (*P*=0.007)** |
| **DeepSurv** | **6.28 [5.30-7.44]\***<br>**(*P*=0.014)** | **57.02 [51.94-61.00]\* (*P*=0.027)** | 91.08 [89.67-92.32] (*P*=0.085) |

## 1.4 Discussion

The progression from traditional statistics to machine learning to deep neural networks continue a trend of identifying ever more complex networks of features and maximizing the value that each feature has in contributing to building of a model. The current study proposes a role for deep neural networks to augment current survival analysis pipelines. DeepSurv, which is a COXPH model that builds on the nonlinear aspect of deep neural networks [13], provides a more robust time-to-event prediction of both mortality and a composite of cardiovascular outcomes. Moreover, this study is one of the first to provide an in-depth examination of the additive value of cardiovascular biomarkers in machine and deep learning survival networks. While we see incremental improvements with the addition of features, the DeepSurv algorithm suggests that traditional risk factors alone are sufficient to provide significant improvement over routine statistical approaches, such as COXPH, and that addition of other features outlined in this study may not be necessary.

Because this is a prospective, survival network spanning 16-years, age discriminates well across both MAE and mortality, likely limiting the impact of other features, such as CAC. Nevertheless, we continue to confirm small, yet significant improvement in model performance with the addition of coronary artery calcium (CAC) confirming previous observations regarding improvements of AUC [23] and NRI [8, 12, 24] with CAC. Examining the MESA cohort, Folsom et al. revealed an increase in the AUC when transitioning from traditional risk factors alone to risk factors including CAC for cardiovascular disease (0.772 vs. 0.811) and coronary heart disease (0.771 vs. 0.824) [23]. In the current study, prediction of MAE for traditional risk factors alone (0.82) and those including CAC (0.85) show a similar progression in improvement. Though, overall, the deep learning algorithms presented in this study are capable of achieving much higher levels of prediction compared to standard statistical approaches. While both the current study and previous research have shown improvements with the addition of CAC, the data presented here provides one of the first examples of how features can be used progressively in machine learning driven survival networks, with the findings indicating modest, at best, improvements with the addition of features beyond traditional risk factors. This concept holds true for not only DeepSurv, but also the other machine and deep learning models tested.

While each of the models may perform comparably with the features that most heavily influence the construction of the model, such as participant age and CAC, the machine and deep learning algorithms differ in the utility they can derive from other less “important” features. DeepSurv outperformed the other models across all features, but DeepSurv’s ability to rely on the traditional risk factors within the feature sets may be its most important attribute. DeepSurv, as hyperparameter optimized in this study, can provide a robust risk score criteria on traditional risk factors alone, significantly improving the classification of true positives. Both NRI and RR highlight the ability of DeepSurv to augment ASCVD prediction and provide a generalizable criterion for risk stratification of patients. This suggests that while all the models show incremental improvement with the addition of features, DeepSurv has the capacity to establish a higher baseline AUC, with a better predictive algorithm for traditional risk factors.

In examining the MESA data, this is the first study to examine both support vector machines (lSVM) as well as deep learning algorithms (nMTLR and DeepSurv) in cardiovascular event prediction for survival analysis. The lSVM provided the second highest prediction scores for both classification of mortality and MAE, acting in a similar fashion to neural networks. SVM functions by employing hyperplanes and n-dimensional spaces, similar to the hidden layers in neural networks [25]. One of the major differences is that SVM work in a linear fashion, whereas neural networks are non-linear algorithms. The strengths of both the lSVM and DeepSurv models presented here likely come from the multi-dimensional structuring of these models, improving their predictive capacity. It is important to note that while DeepSurv provided the best prediction in this study, further hyperparameter optimization of DeepSurv or parameter tuning of the other machine learning models, including lSVM, could increase their predictive power.

The current study is limited by a lack of true optimization for all of the machine and deep learning models. DeepSurv was the only algorithm optimized for the current studies, whereas the other machine and deep learning algorithms were used with the “out-of-the-box” functions. This warrants a disclaimer that improvement in prediction of these algorithms can likely be made, even in the case of DeepSurv, which could undergo further hyperparameter optimization. Though machine and deep learning provide progressive leaps for clinical medicine, unlike traditional statistical methods, parameter optimization for each dataset is needed to unlock the algorithm’s complete potential.

## 1.5 Conclusions

In the MESA cohort, we provide an examination of machine and deep learning approaches applied to survival analysis. We highlight the ability of deep neural networks to modestly increase the predictive capacity of our endpoints, including cardiovascular outcomes and death. DeepSurv has the ability to gain additional information from traditional risk factors within the model and leverage its ability to form new, non-linear connections, providing unique associations within the dataset. Ultimately, these machine and deep learning models can be implemented as risk scores, creating generalizable models that can inform clinical prognoses.

## Data Availability

Computations were performed in Python 3.7, all code is provided: https://github.com/qahathaway/MESA.

## 1.6 Acknowledgements

MESA and the MESA SHARe project are conducted and supported by the National Heart, Lung, and Blood Institute (NHLBI) in collaboration with MESA investigators. Support for MESA is provided by contracts N01-HC95159, N01-HC-95160, N01-HC-95161, N01-HC-95162, N01-HC-95163, N01-HC-95164, N01-HC-95165, N01-HC95166, N01-HC-95167, N01-HC-95168, N01-HC-95169 and CTSA UL1-RR-024156. This manuscript was not prepared in collaboration with MESA investigators and does not necessarily reflect the opinions or views of MESA, or the NHLBI.

## 1.9 Author Contributions

Conceptualization (QAH, NY, MJB, PPS, IZ); Data curation (QAH, NY, PPS, IZ); Formal analysis (QAH, NY); Funding acquisition (QAH, PPS); Investigation (QAH, NY, MJB, PPS, IZ); Methodology (QAH, NY, MJB, PPS, IZ); Project administration (PPS, IZ); Resources (PPS); Software (QAH, NY, PPS); Supervision (PPS, IZ); Validation (QAH, NY, MJB, PPS, IZ); Visualization (QAH, NY); Roles/Writing - original draft (QAH, NY, IZ); Writing - review & editing (MJB, PPS).

---

**Supplemental Figure 1.**
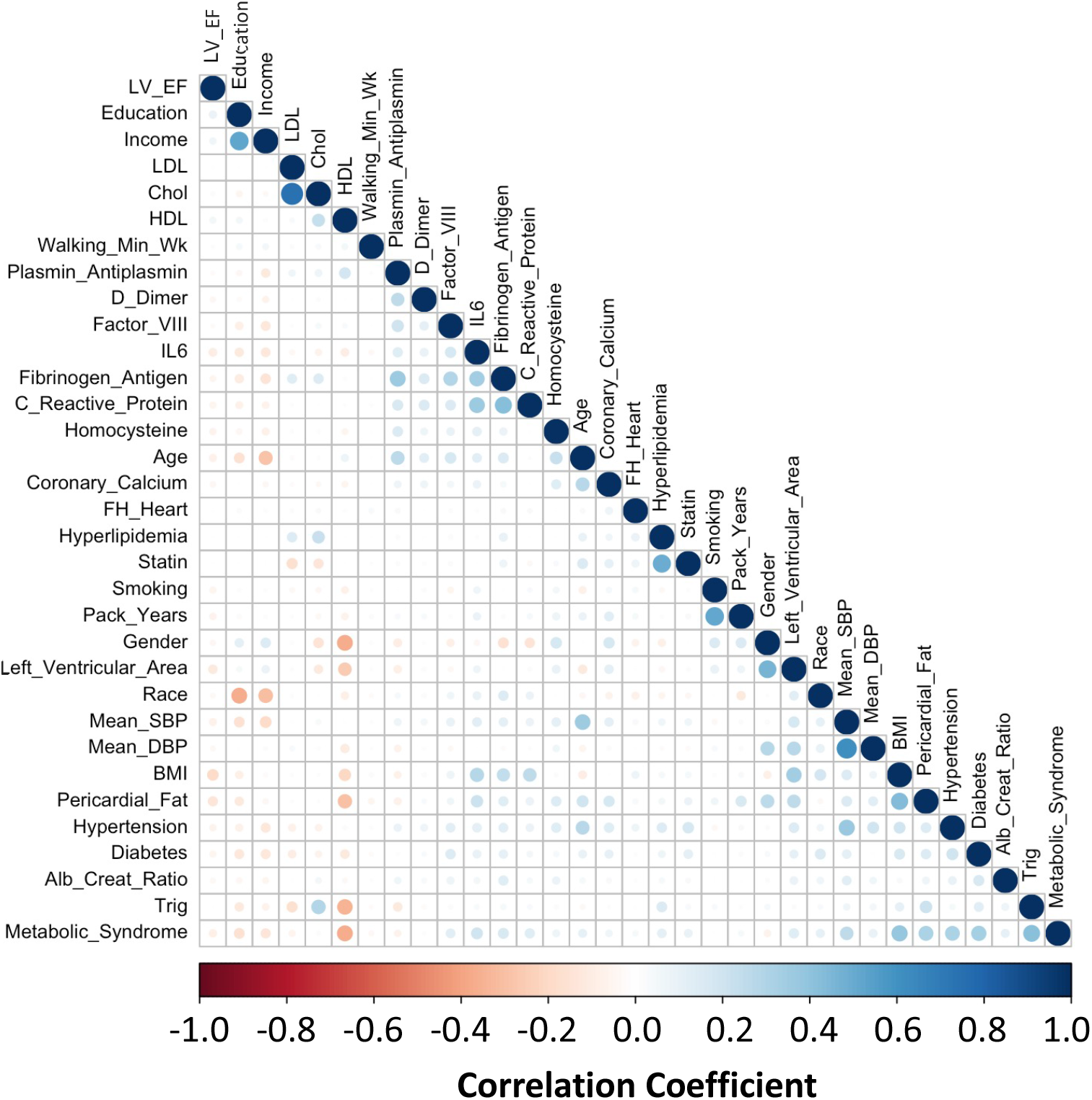

**Supplemental Figure 2.**
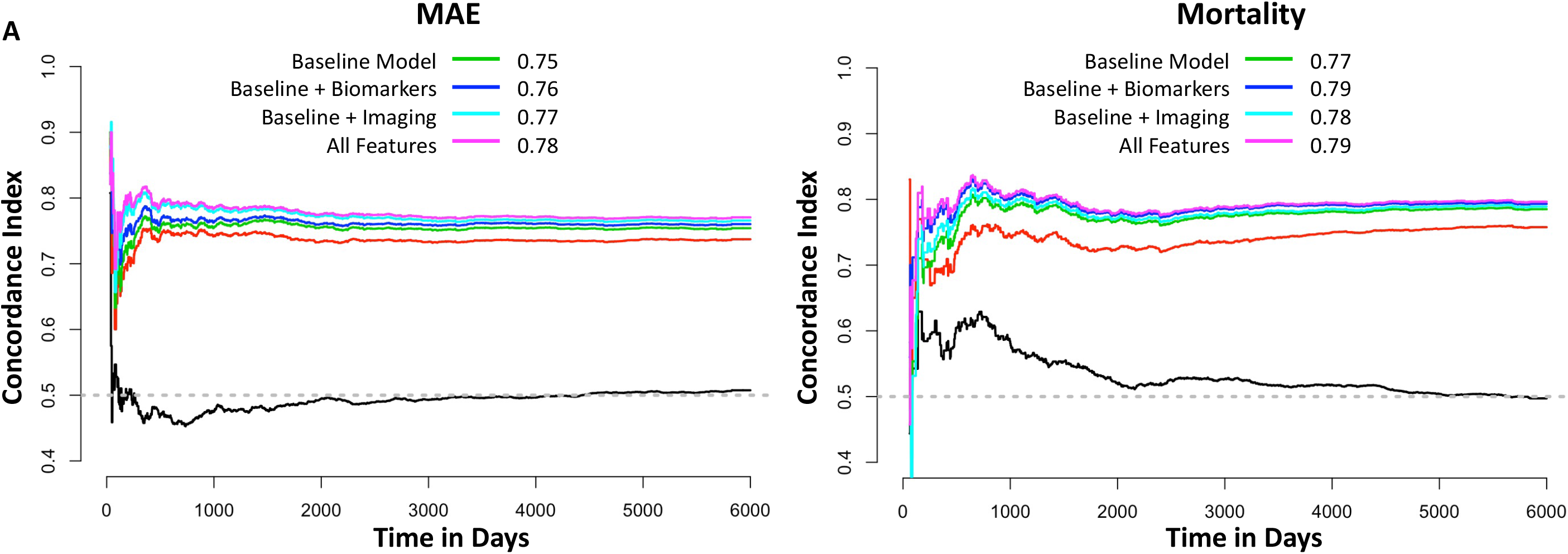

**Supplemental Figure 3.**
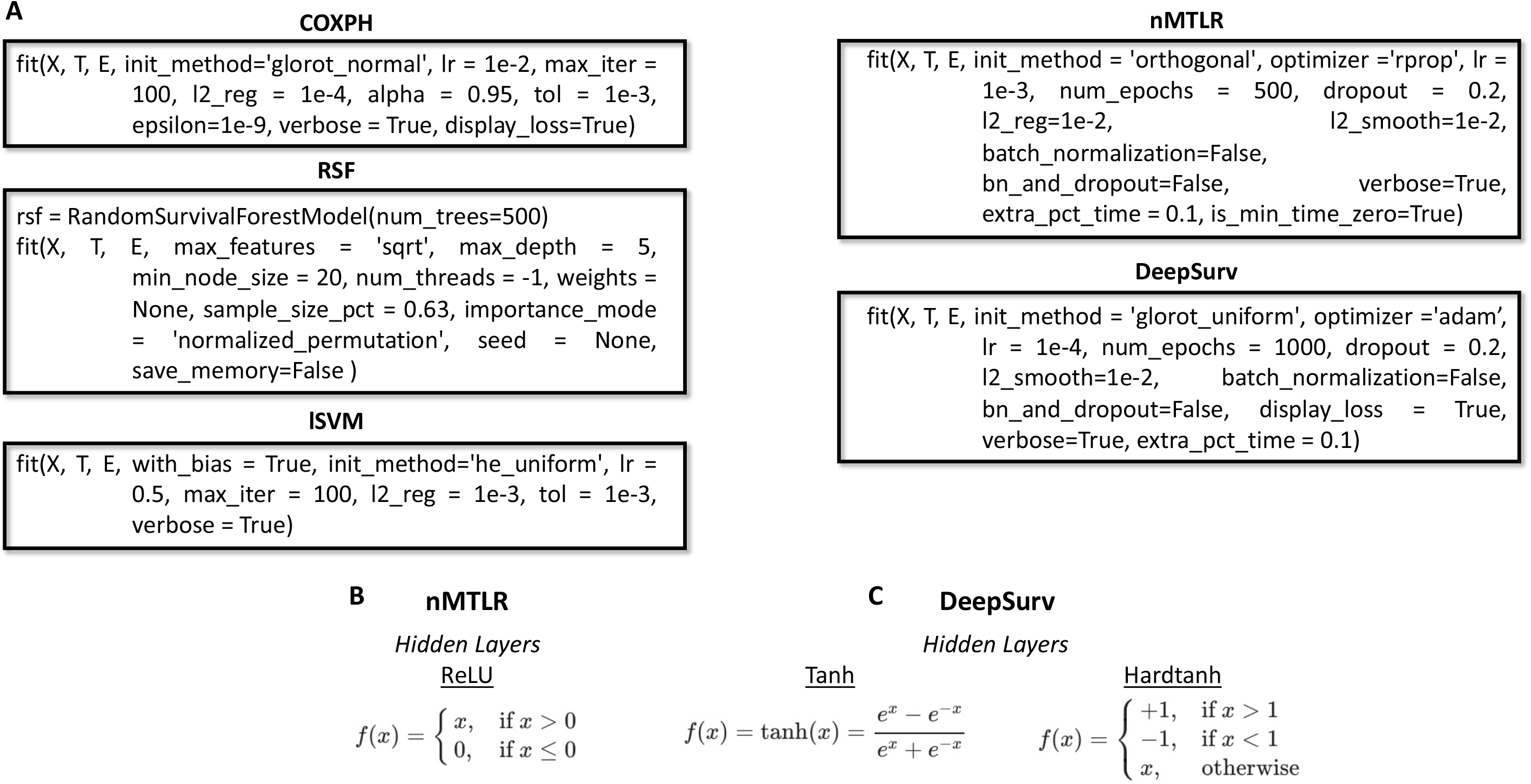

**Supplemental Figure 4.**
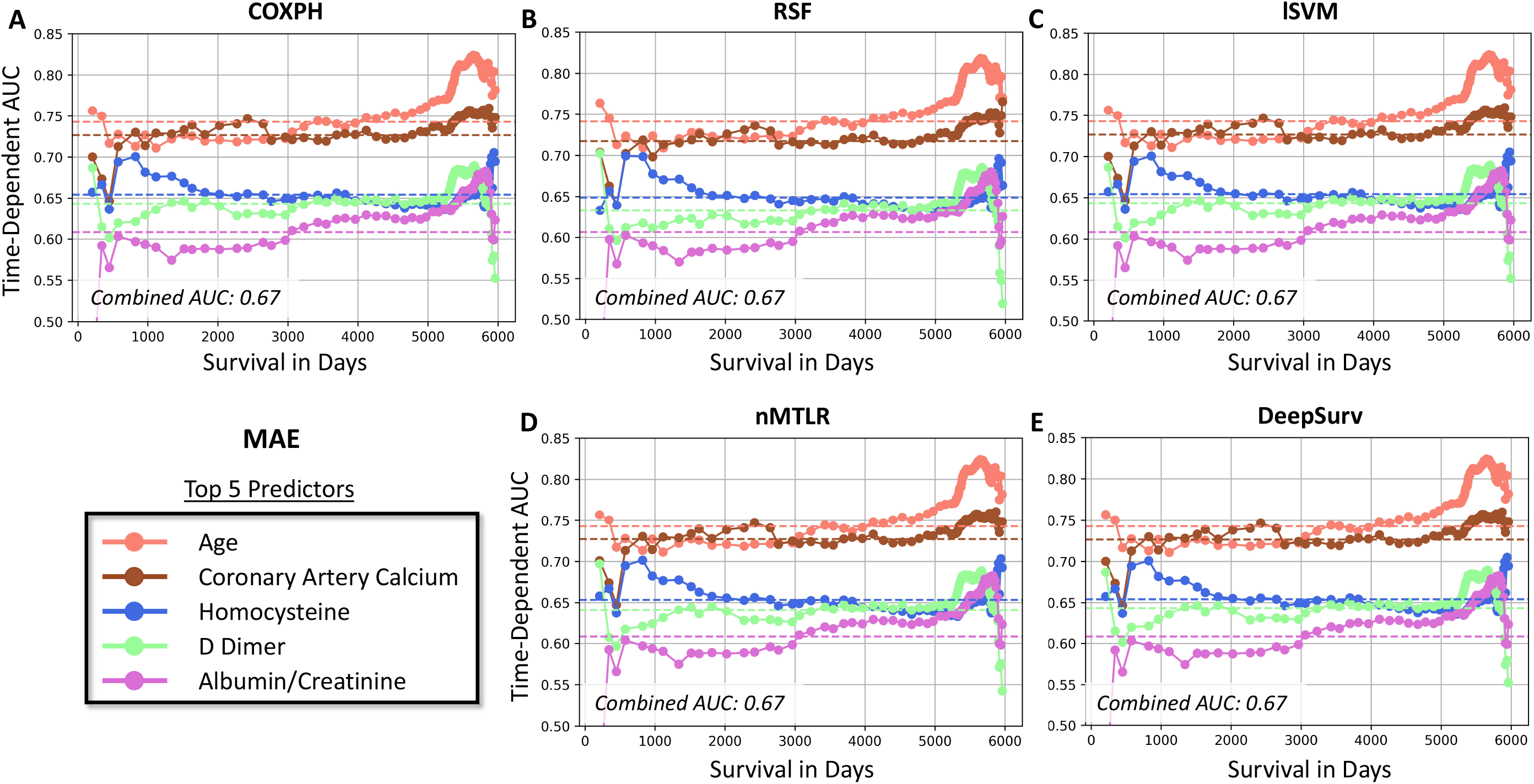

**Supplemental Figure 5.**
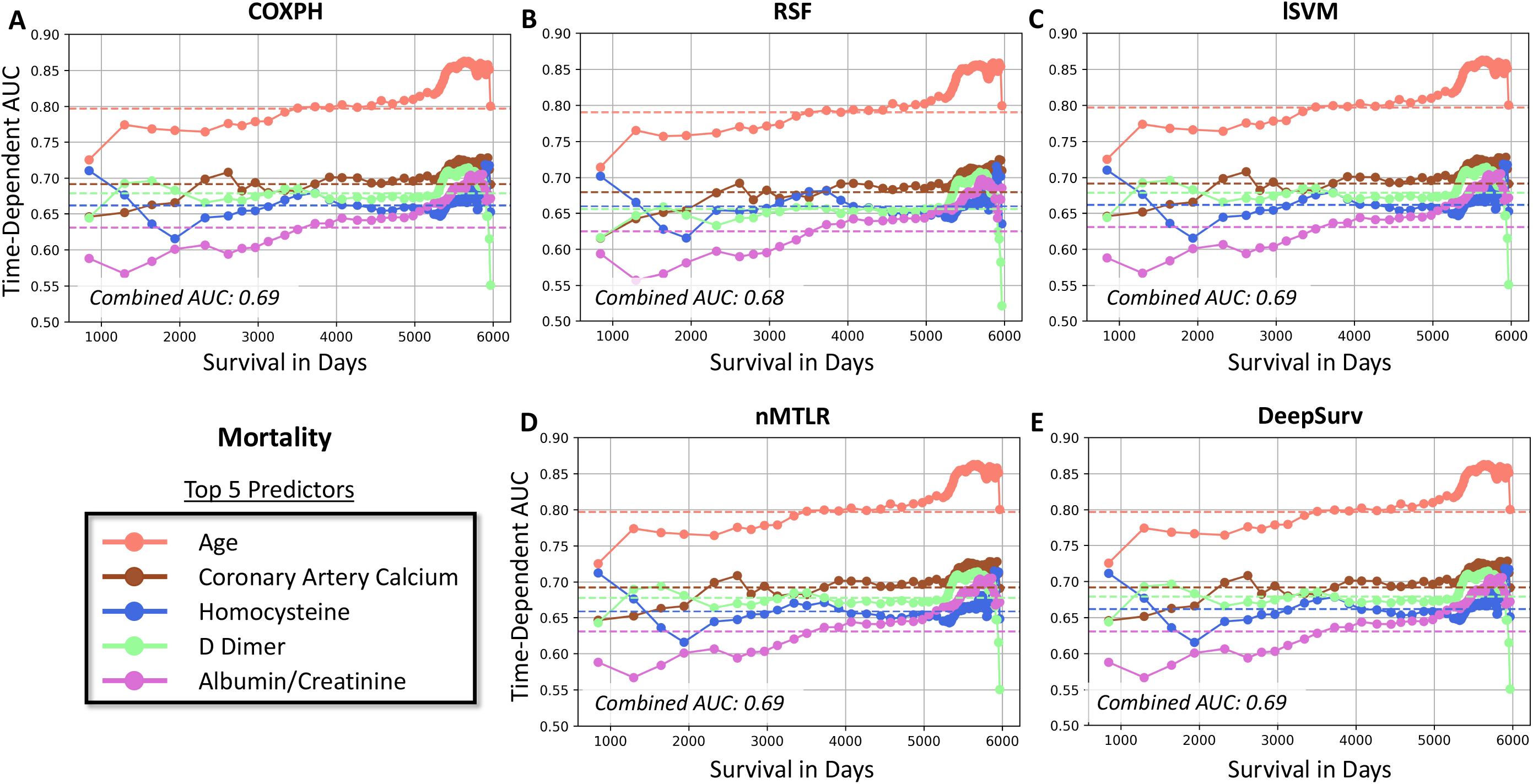

**Supplemental Figure 6.**
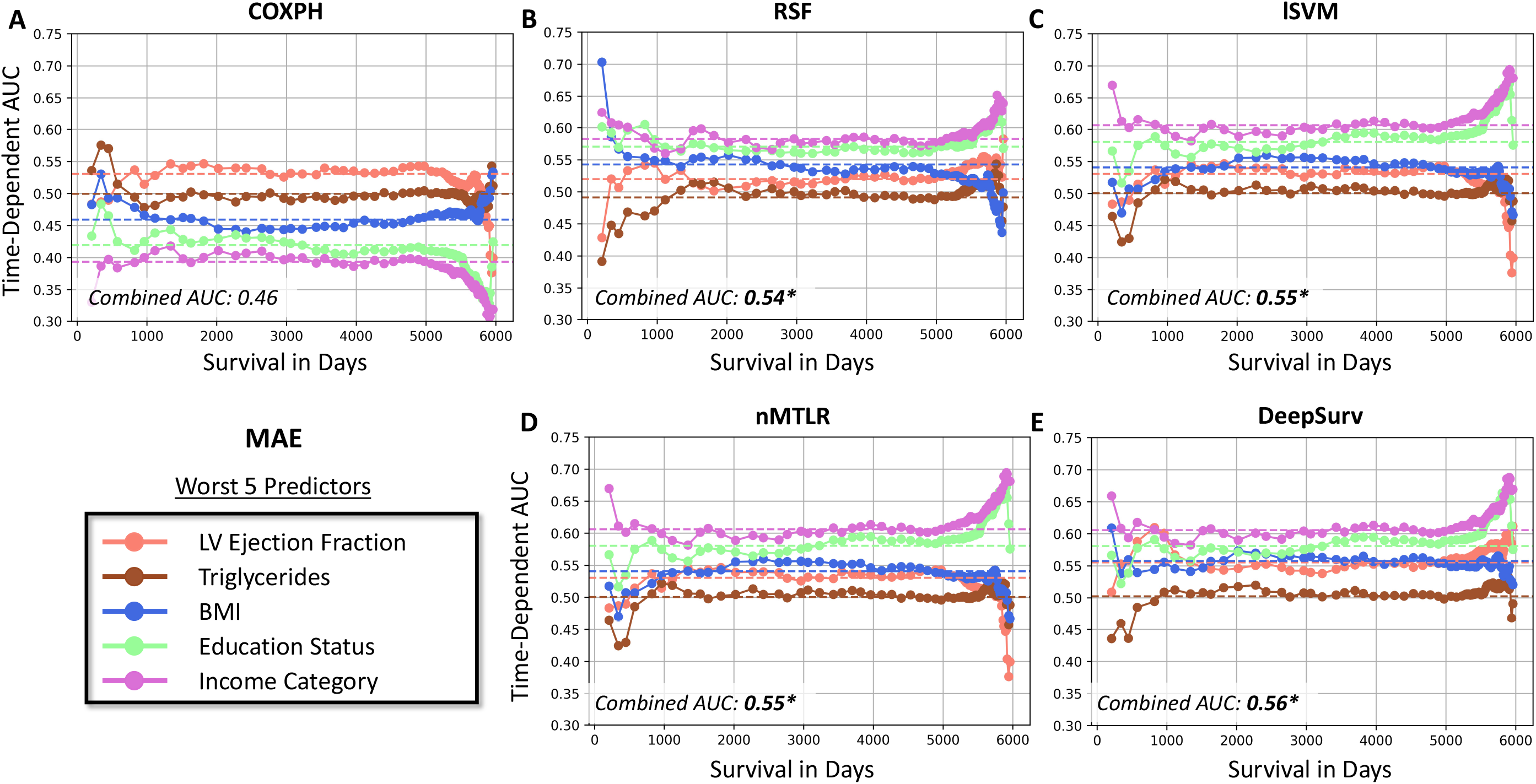

**Supplemental Figure 7.**
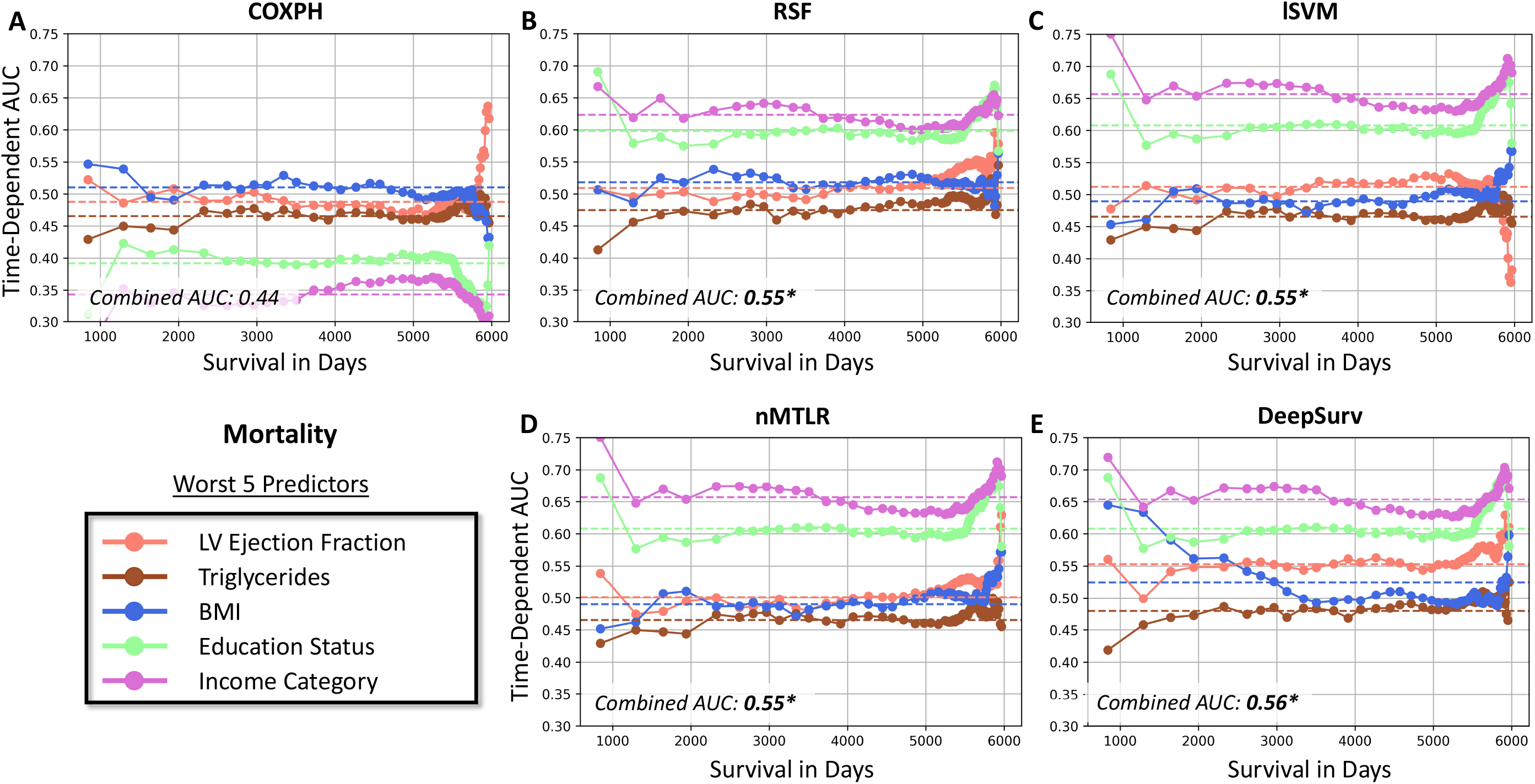

