## Supplemental Methods, Tables, and Figure Legends for "Deep Neural Survival Networks for Cardiovascular Risk Prediction: The Multi-Ethnic Study of Atherosclerosis (MESA)"

***Running Title:*** *Deep Neural Survival Networks: MESA*

**Corresponding Author**

Partho P. Sengupta, MD, DM

PO Box 8003

WVU Heart & Vascular Institute,

1 Medical Center Drive, Morgantown, WV 26506.

**Supplemental Materials and Methods**

*The Multi-Ethnic Study of Atherosclerosis (MESA) Cohort*

For complete information regarding the study design, objectives, approval, and participant consent, refer to Bild et al. [1]. Each field center was approved for the outlined study protocol by institutional review boards, with all participants giving informed consent [1]. For a more thorough understanding of how features were acquired from participants, refer to Ambale-Venkatesh et al. [2]. We included all-cause mortality because the etiology of death is, usually, multifactorial and cardiac contribution may be underappreciated. Moreover, the recorded cause of death, if not observed in a clinical environment, could be inaccurate and the inclusion of all-cause mortality in the composite outcome reduces the risk of bias by competing risks. We also performed analyses using all-cause mortality as an independent end point.

*Machine and Deep Learning Models*

All five models were employed through the algorithms in the Pysurvival toolbox [3], with modifications. Briefly, all models were executed with the standard configurations outlined in the Pysurvival toolbox, with the exception of the RSF and DeepSurv models. The number of trees in the RSF model was increased to 500 and the DeepSurv model added an additional hidden layer, which were determined through hyperparameter optimization **(Supplemental Figure 3)**. Each of the five models were assessed independently with the following feature compositions: Baseline Model, Biomarkers, and Imaging. Additionally, each of the models were assessed with all of the features unique to the Baseline Model, Biomarkers, and Imaging datasets, identified as All Features.

*Evaluation of Machine and Deep Learning Models*

Assessments of each model were performed using traditional evaluation markers, such as assessing the number of concordant pairs of predicted and observed outcomes (Harrell’s concordance index) and the mean squared difference between the predicted and observed probabilities of an outcome (integrated brier score) [4]. Because the dataset is highly right-censored **(Figure 1B)**, the inverse probability of censoring weights, or concordance index IPCW [5], was used to accurately depict the concordance index for the survival analyses. Additionally, the time-dependent AUC (cumulative/dynamic) [6] was used to determine the sensitivity and specificity of prediction at given intervals along the survival curve. The concordance index and the integrated brier score were calculated using the Pysurvival toolbox [3]. The concordance index IPCW and time-dependent AUC were processed through scikit-survival 0.12.0 [7] and scikit-learn 0.23.1.

To determine the clinical importance of machine and deep learning algorithms, net reclassification improvement (NRI) and relative risk (RR) were evaluated. The R package NRIcens 1.6 was used to calculate the non-categorical NRI with the definition from Pencina et al. [8, 9], using dependencies from the R package survival 3.2. [10]. This definition is the probability of $\left( \frac{(Cases correctly reclassified to a higher risk category}{Event}- \frac{Cases incorrectly reclassified to a lower risk category}{Event} \right)+\left( \frac{(Controls correctly reclassified to a lower risk category}{No Event}- \frac{Controls incorrectly reclassified to a higher risk category}{No Event} \right)$. This interpretation of the NRI allows for a range that encompasses a perfect incorrect reclassification (-200%) to a perfect reclassification (200%). The non-categorical NRI was assessed at 15 years (5,475 days), with cases defined as those with an event before 15 years and controls defined as participants censored after 15 years. A categorical NRI was used to assess reclassification at risk intervals through PredictABEL 1.2-4 [11]. The Pooled Cohort Equation (PCE) estimated 10-year risk for atherosclerotic cardiovascular disease (ASCVD) was implemented to define categorical cutoffs equated to 5% (0.27), 7.5% (0.36), and 20% (0.78) risk of event.

All evaluations were implemented on the external validation set, containing the participants from field centers 6 and 7. The concordance index and concordance index IPCW were calculated using bins of 50 different time points within the external validation set, for a total of 28 observations. The integrated brier score and cumulative/dynamic time-depend AUC were also calculated at 28 observations within the external validation set. Risk scores were generated using the Pysurvival toolbox function *predict_risk*. Outliers in the risk score were determined by the quartile equation: $Q3+(1.5*IQR)$, where Q3 is the third quartile and IQR is the interquartile range. Numbers exceeding the quartile equation were set equal to the quartile equation as the defined upper limit. Risk scores were further normalized, using the mean and standard deviation of the data. Computations were performed in Python 3.7, all code is provided: <https://github.com/qahathaway/MESA>.

**Tables and Table Legends:**

**Supplemental Table 1: Time-Dependent AUC – Race/Ethnicity**

| **MAE** | | | | |
| --- | --- | --- | --- | --- |
| **Models/Risk Scores** | **White/Caucasian** | **Chinese American** | **Black/African American** | **Hispanic** |
| **COXPH – Baseline Model** | 0.80 [0.78-0.81] | 0.77 [0.74-0.81] | 0.74 [0.74-0.75] | 0.78 [0.77-0.79] |
| **DeepSurv – Baseline Model** | **0.82 [0.81-0.83]* (*P*=0.005)** | **0.82 [0.79-0.85]* (*P*=0.025)** | **0.77 [0.76-0.78]*** (***P*≤0.001)** | **0.81 [0.80-0.82]*** (***P*≤0.001)** |
| **COXPH – All Features** | 0.80 [0.79-0.81] | 0.71 [0.69-0.73] | 0.77 [0.76-0.78] | 0.75 [0.74-0.75] |
| **DeepSurv – All Features** | 0.81 [0.80-0.82] (*P*=0.324) | **0.83 [0.79-0.86]* (*P*≤0.001)** | **0.79 [0.77-0.80]* (*P*=0.018)** | **0.81 [0.80-0.82]* (*P*≤0.001)** |
| **Mortality** | | | | |
| **Models/Risk Scores** | **White/Caucasian** | **Chinese American** | **Black/African American** | **Hispanic** |
| **COXPH – Baseline Model** | 0.83 [0.82-0.84] | 0.70 [0.68-0.72] | 0.75 [0.74-0.76] | 0.81 [0.80-0.83] |
| **DeepSurv – Baseline Model** | **0.85 [0.84-0.86]* (*P*=0.034)** | **0.84 [0.81-0.86]* (*P*≤0.001)** | **0.81 [0.80-0.82]*** (***P*≤0.001)** | **0.88 [0.87-0.90]*** (***P*≤0.001)** |
| **COXPH – All Features** | 0.80 [0.79-0.81] | 0.79 [0.78-0.81] | 0.80 [0.78-0.81] | 0.85 [0.84-0.87] |
| **DeepSurv – All Features** | **0.84 [0.83-0.85]* (*P*≤0.001)** | **0.83 [0.81-0.84]* (*P*≤0.001)** | **0.84 [0.83-0.85]* (*P*≤0.001)** | 0.85 [0.84-0.86] (P=0.731) |

**Supplemental Table 1:** Cumulative/dynamic time-dependent AUCs for race/ethnicity groups for DeepSurv utilizing the Baseline Model features. Data are statistically significant with a *P* ≤ 0.05 when denoted by a “*****”. Data are represented as the mean and the 95% confidence interval. MAE = major adverse events, including all-cause mortality and cardiovascular outcomes, Baseline Model = traditional risk factors, All Features = traditional risk factors, inflammatory biomarkers, and left ventricular structure and function, COXPH = Cox Proportional Hazards Regression Model, DeepSurv = Non-Linear Cox Proportional Hazards Deep Neural Network.

**Supplemental Table 2: Concordance Index**

| **MAE** | | | | |
| --- | --- | --- | --- | --- |
| **Features** | **Baseline Model** | **Baseline + Biomarkers** | **Baseline + Imaging** | **All Features** |
| **COXPH [95% CI]** | 0.74 [0.72-0.76] | 0.73 [0.71-0.75] | 0.75 [0.72-0.77] | 0.75 [0.71-0.78] |
| **nMTLR [95% CI]** | 0.74 [0.72-0.76] (*P*=0.997) | 0.74 [0.72-0.77] | 0.76 [0.74-0.78] | 0.77 [0.75-0.78] (*P*=0.302) |
| **RSF [95% CI]** | 0.75 [0.73-0.77] (*P*=0.927) | 0.76 [0.74-0.78] | 0.79 [0.77-0.80] | **0.78 [0.77-0.80]* (*P*=0.013)** |
| **lSVM [95% CI]** | 0.77 [0.75-0.79] (*P*=0.162) | 0.77 [0.75-0.79] | 0.78 [0.76-0.80] | **0.78 [0.76-0.80]* (*P*=0.047)** |
| **DeepSurv [95% CI]** | **0.78 [0.76-0.80]* (*P*=0.039)** | 0.78 [0.76-0.80] | 0.79 [0.77-0.81] | **0.79 [0.78-0.81]* (*P*=0.004)** |
| **Mortality** | | | | |
| **COXPH [95% CI]** | 0.78 [0.75-0.80] | 0.78 [0.77-0.80] | 0.79 [0.76-0.82] | 0.79 [0.78-0.81] |
| **nMTLR [95% CI]** | 0.80 [0.78-0.83] (*P*=0.392) | 0.76 [0.72-0.79] | 0.80 [0.78-0.83] | 0.78 [0.76-0.80] (*P*=0.647) |
| **RSF [95% CI]** | 0.80 [0.78-0.82] (*P*=0.485) | 0.81 [0.79-0.83] | 0.81 [0.80-0.83] | 0.81 [0.79-0.83] (*P*=0.663) |
| **lSVM [95% CI]** | 0.81 [0.79-0.83] (*P*=0.165) | 0.82 [0.80-0.84] | 0.82 [0.80-0.84] | 0.82 [0.80-0.85] (*P*=0.120) |
| **DeepSurv [95% CI]** | **0.83 [0.81-0.85]* (*P*=0.012)** | 0.83 [0.80-0.85] | 0.83 [0.81-0.85] | **0.84 [0.81-0.86]* (*P*=0.009)** |

**Supplemental Table 2:** Concordance index for the COXPH, RSF, lSVM, nMTLR, and DeepSurv models. Data are statistically significant with a *P* ≤ 0.05 when denoted by a “*****”. Data are represented as the mean and the 95% confidence interval. MAE = major adverse events, including all-cause mortality and cardiovascular outcomes, Baseline Model = traditional risk factors, Biomarkers = inflammatory biomarkers, Imaging = left ventricular structure and function, All Features = traditional risk factors, inflammatory biomarkers, and left ventricular structure and function, COXPH = Cox Proportional Hazards Regression Model, RSF = Random Survival Forest, lSVM = Linear Support Vector Machines, nMTLR = Neural Multi-Task Logistic Regression, DeepSurv = Non-Linear Cox Proportional Hazards Deep Neural Network.

**Supplemental Table 3: Concordance Index IPCW**

| **MAE** | | | | |
| --- | --- | --- | --- | --- |
| **Features** | **Baseline Model** | **Baseline + Biomarkers** | **Baseline + Imaging** | **All Features** |
| **COXPH [95% CI]** | 0.74 [0.72-0.76] | 0.73 [0.71-0.75] | 0.76 [0.73-0.78] | 0.75 [0.73-0.77] |
| **nMTLR [95% CI]** | 0.73 [0.71-0.76] (*P*=0.989) | 0.74 [0.72-0.76] | 0.76 [0.74-0.78] | 0.77 [0.75-0.78] (*P*=0.631) |
| **RSF [95% CI]** | 0.75 [0.73-0.77] (*P*=0.866) | 0.75 [0.73-0.78] | 0.78 [0.76-0.80] | 0.78 [0.77-0.80] (*P*=0.065) |
| **lSVM [95% CI]** | 0.77 [0.75-0.79] (*P*=0.143) | 0.77 [0.75-0.79] | 0.78 [0.76-0.80] | 0.78 [0.76-0.80] (*P*=0.152) |
| **DeepSurv [95% CI]** | **0.77 [0.75-0.79]* (*P*=0.046)** | 0.78 [0.76-0.80] | 0.79 [0.77-0.81] | **0.79 [0.77-0.81]* (*P*=0.030)** |
| **Mortality** | | | | |
| **COXPH [95% CI]** | 0.78 [0.76-0.80] | 0.78 [0.77-0.80] | 0.79 [0.76-0.81] | 0.79 [0.77-0.81] |
| **nMTLR [95% CI]** | 0.80 [0.78-0.83] (*P*=0.491) | 0.77 [0.74-0.79] | 0.80 [0.77-0.83] | 0.78 [0.75-0.80] (*P*=0.739) |
| **RSF [95% CI]** | 0.80 [0.77-0.83] (*P*=0.508) | 0.80 [0.78-0.83] | 0.81 [0.79-0.84] | 0.81 [0.78-0.83] (*P*=0.619) |
| **lSVM [95% CI]** | 0.82 [0.79-0.84] (*P*=0.094) | 0.82 [0.80-0.84] | 0.82 [0.80-0.83] | 0.82 [0.80-0.83] (*P*=0.145) |
| **DeepSurv [95% CI]** | **0.83 [0.80-0.85]* (*P*=0.018)** | 0.83 [0.80-0.85] | 0.81 [0.81-0.85] | **0.84 [0.81-0.86]* (*P*=0.012)** |

**Supplemental Table 3:** Concordance index with inverse probability of censoring weights (IPCW) for the COXPH, RSF, lSVM, nMTLR, and DeepSurv models. Data are statistically significant with a *P* ≤ 0.05 when denoted by a “*****”. Data are represented as the mean and the 95% confidence interval. MAE = major adverse events, including all-cause mortality and cardiovascular outcomes, Baseline Model = traditional risk factors, Biomarkers = inflammatory biomarkers, Imaging = left ventricular structure and function, All Features = traditional risk factors, inflammatory biomarkers, and left ventricular structure and function, COXPH = Cox Proportional Hazards Regression Model, RSF = Random Survival Forest, lSVM = Linear Support Vector Machines, nMTLR = Neural Multi-Task Logistic Regression, DeepSurv = Non-Linear Cox Proportional Hazards Deep Neural Network.

**Supplemental Table 4: Integrated Brier Score (IBS)**

| **MAE** | | | | |
| --- | --- | --- | --- | --- |
| **Features** | **Baseline Model** | **Baseline + Biomarkers** | **Baseline + Imaging** | **All Features** |
| **COXPH [95% CI]** | 0.09 [0.09-0.09] | 0.09 [0.09-0.09] | 0.09 [0.09-0.09] | 0.09 [0.09-0.09] |
| **nMTLR [95% CI]** | 0.10 [0.09-0.10] (*P*=0.3438) | 0.10 [0.09-0.11] | 0.10 [0.09-0.11] | 0.10 [0.09-0.11] (*P*=0.097) |
| **RSF [95% CI]** | **0.11 [0.11-0.11]* (*P*≤0.001)** | 0.11 [0.11-0.11] | 0.10 [0.10-0.11] | **0.10 [0.10-0.11]* (*P*≤0.001)** |
| **lSVM [95% CI]** | *N/A* | *N/A* | *N/A* | *N/A* |
| **DeepSurv [95% CI]** | 0.09 [0.08-0.09] (*P*=0.251) | 0.09 [0.08-0.09] | 0.08 [0.08-0.09] | **0.08 [0.08-0.09]* (*P*=0.005)** |
| **Mortality** | | | | |
| **COXPH [95% CI]** | 0.07 [0.07-0.07] | 0.07 [0.07-0.07] | 0.07 [0.06-0.07] | 0.06 [0.06-0.07] |
| **nMTLR [95% CI]** | **0.06 [0.05-0.07]* (*P*=0.014)** | 0.06 [0.05-0.07] | 0.06 [0.05-0.07] | 0.06 [0.06-0.07] (*P*=0.929) |
| **RSF [95% CI]** | **0.08 [0.08-0.09]* (*P*≤0.001)** | 0.08 [0.08-0.09] | 0.08 [0.08-0.08] | **0.08 [0.08-0.08]* (*P*≤0.001)** |
| **lSVM [95% CI]** | *N/A* | *N/A* | *N/A* | *N/A* |
| **DeepSurv [95% CI]** | **0.06 [0.06-0.07]* (*P*=0.008)** | 0.06 [0.06-0.06] | 0.06 [0.06-0.07] | 0.06 [0.06-0.06] (*P*=0.309) |

**Supplemental Table 4:** Integrated brier score for the COXPH, RSF, nMTLR, and DeepSurv models. Integrated brier score cannot be calculated for lSVM. Data are statistically significant with a *P* ≤ 0.05 when denoted by a “*****”. Data are represented as the mean and the 95% confidence interval. MAE = major adverse events, including all-cause mortality and cardiovascular outcomes, Baseline Model = traditional risk factors, Biomarkers = inflammatory biomarkers, Imaging = left ventricular structure and function, All Features = traditional risk factors, inflammatory biomarkers, and left ventricular structure and function, COXPH = Cox Proportional Hazards Regression Model, RSF = Random Survival Forest, lSVM = Linear Support Vector Machines, nMTLR = Neural Multi-Task Logistic Regression, DeepSurv = Non-Linear Cox Proportional Hazards Deep Neural Network.

**Supplemental Table 5: Reclassification – All Features**

|  | **Non-Categorical NRI (%) [95% CI]** | **Cases – Increased Risk Category (%) [95% CI]** | **Controls – Decreased Risk Category (%) [95% CI]** |
| --- | --- | --- | --- |
| **MAE** | | | |
| **Risk Scores** | **All Features [95% CI]** | | |
| **COXPH** | 0.00 | 50.00 | 50.00 |
| **nMTLR** | **44.61 [35.55-53.68]* (*P*≤0.05)** | 64.93 [62.18-69.60] | 57.38 [54.28-61.01] |
| **RSF** | **57.02 [44.97-65.88]* (*P*≤0.05)** | 65.54 [61.26-68.53] | 62.97 [60.33-64.92] |
| **lSVM** | **58.36 [51.52-68.15]* (*P*≤0.05)** | 62.58 [58.46-66.29] | 66.60 [63.30-69.72] |
| **DeepSurv** | **63.88 [56.71-72.31]* (*P*≤0.05)** | 70.70 [67.91-74.94] | 61.24 [58.79-64.24] |
| **Mortality** | | | |
| **Risk Scores** | **All Features [95% CI]** | | |
| **COXPH** | 0.00 | 50.00 | 50.00 |
| **nMTLR** | **31.76 [21.89-42.11]* (*P*≤0.05)** | 63.71 [59.27-68.28] | 52.17 [48.83-55.26] |
| **RSF** | **43.44 [32.59-54.11]* (*P*≤0.05)** | 62.30 [56.34-67.56] | 59.42 [56.95-61.55] |
| **lSVM** | **58.32 [39.93-56.32]* (*P*≤0.05)** | 64.54 [59.71-67.42] | 64.62 [59.71-69.35] |
| **DeepSurv** | **61.30 [50.84-75.17]* (*P*≤0.05)** | 69.27 [65.06-74.39] | 61.38 [59.77-63.38] |

**Supplemental Table 5:** Non-categorical net reclassification improvement (NRI) and the percentage of participants moving up and down risk categories for the COXPH, RSF, lSVM, nMTLR, and DeepSurv models. Analyses performed on All Features. Data are statistically significant with a *P* ≤ 0.05 when denoted by a “*****”. Data are represented as the mean and the 95% confidence interval. MAE = major adverse events, including all-cause mortality and cardiovascular outcomes, Cases = participant with an event, Controls = participants without an event, All Features = traditional risk factors, inflammatory biomarkers, and left ventricular structure and function, COXPH = Cox Proportional Hazards Regression Model, RSF = Random Survival Forest, lSVM = Linear Support Vector Machines, nMTLR = Neural Multi-Task Logistic Regression, DeepSurv = Non-Linear Cox Proportional Hazards Deep Neural Network.

**Supplemental Table 6: Confusion Matrix – Categorical NRI**

|  | **MAE - Baseline** | | | | |  | **MAE – All Features** | | | | |
| --- | --- | --- | --- | --- | --- | --- | --- | --- | --- | --- | --- |
|  | **U – [0,0.27)** | **U – [0.27,0.36)** | **U – [0.36,0.78)** | **U – [0.78,1]** | **U – reclassified** |  | **U – [0,0.27)** | **U – [0.27,0.36)** | **U – [0.36,0.78)** | **U – [0.78,1]** | **U – reclassified** |
| **I – [0,0.27)** | 666 | 107 | 100 | 9 | 24% |  | 558 | 146 | 113 | 23 | 34% |
| **I – [0.27,0.36)** | 127 | 79 | 136 | 21 | 78% |  | 145 | 76 | 131 | 40 | 81% |
| **I – [0.36,0.78)** | 49 | 95 | 238 | 128 | 53% |  | 116 | 100 | 198 | 136 | 64% |
| **I – [0.78,1]** | 10 | 26 | 144 | 294 | 38% |  | 18 | 36 | 131 | 262 | 41% |
|  | **Mortality - Baseline** | | | | |  | **Mortality – All Features** | | | | |
| **I – [0,0.27)** | 760 | 107 | 30 | 3 | 16% |  | 698 | 110 | 64 | 6 | 21% |
| **I – [0.27,0.36)** | 85 | 132 | 151 | 6 | 65% |  | 146 | 112 | 146 | 19 | 74% |
| **I – [0.36,0.78)** | 19 | 64 | 316 | 103 | 37% |  | 49 | 89 | 217 | 109 | 53% |
| **I – [0.78,1]** | 4 | 4 | 100 | 345 | 24% |  | 5 | 12 | 123 | 324 | 30% |

**Supplemental Table 6:** Categorical net reclassification improvement (NRI) for low (0-5% or 0.0-0.27), borderline (5%-7.5%, or 0.27-0.36), intermediate (7.5%-20% or 0.36-0.78), and high (>20% or 0.78-1) risk participants. COXPH was used as the initial model (I) and DeepSurv as the updated model (U). Analyses performed on the Baseline Model and All Features. Data are statistically significant with a *P* ≤ 0.05 when denoted by a “*****”. Data are represented as the mean and the 95% confidence interval. MAE = major adverse events, including all-cause mortality and cardiovascular outcomes, Baseline Model = traditional risk factors, All Features = traditional risk factors, inflammatory biomarkers, and left ventricular structure and function, I = initial model, U = updated model, COXPH = Cox Proportional Hazards Regression Model, DeepSurv = Non-Linear Cox Proportional Hazards Deep Neural Network.

**Supplemental Table 7: Predicted Risk – All Features**

|  | **Relative Risk [95% CI]** | **Positive Predictive Value (%) [95% CI]** | **Negative Predictive Value (%) [95% CI]** |
| --- | --- | --- | --- |
| **MAE** | | | |
| **Risk Scores** | **All Features [95% CI]** | | |
| **COXPH** | 3.38 [3.08-3.70] | 58.77 [54.17-63.39] | 82.46 [80.52-84.52] |
| **nMTLR** | 3.35 [2.95-3.80] (*P*=1.000) | 59.38 [54.80-63.81] (*P*=0.853) | 82.26 [80.42-83.97] (*P*=1.000) |
| **RSF** | **4.33 [3.81-4.92]* (*P=*0.002)** | **66.25 [61.89-70.35]* (*P=*0.021)** | 84.70 [82.94-86.31] (*P=*0.094) |
| **lSVM** | **4.01 [3.51-4.58]* (*P=*0.038)** | 59.82 [55.71-63.80] (*P*=0.738) | 85.08 [83.29-86.71] (*P=*0.052) |
| **DeepSurv** | **4.28 [3.77-4.85]* (*P=*0.003)** | **66.81 [62.39-70.96]* (*P=*0.013)** | 84.39 [82.62-86.01] (*P*=0.150) |
| **Mortality** | | | |
| **Risk Scores** | **All Features [95% CI]** | | |
| **COXPH** | 4.23 [3.59-4.98] | 46.98 [42.48-51.53] | 88.90 [87.34-90.28] |
| **nMTLR** | 4.39 [3.73-5.16] (*P*=0.752) | 48.55 [43.94-53.17] (*P*=0.635) | 88.95 [87.40-90.32] (*P=*0.962) |
| **RSF** | **6.25 [5.27-7.40]* (*P*≤0.001)** | **55.77 [51.20-60.25]* (*P=*0.008)** | **91.07 [89.65-92.31]* (*P=*0.032)** |
| **lSVM** | **6.58 [5.46-7.92]* (*P*≤0.001)** | 50.09 [46.01-54.16] (*P*=0.320) | **92.38 [91.00-93.56]* (*P*≤0.001)** |
| **DeepSurv** | **7.02 [5.91-8.35]* (*P*≤0.001)** | **58.30 [53.73-62.73]* (*P*≤0.001)** | **91.70 [90.32-92.90]* (*P=*0.005)** |

**Supplemental Table 7:** Relative risk (RR), positive predictive value (PPV), and negative predictive value (NPV) for the COXPH, RSF, lSVM, nMTLR, and DeepSurv models. Analyses performed on All Features. Data are statistically significant with a *P* ≤ 0.05 when denoted by a “*****”. Data are represented as the mean and the 95% confidence interval. MAE = major adverse events, including all-cause mortality and cardiovascular outcomes, All Features = traditional risk factors, inflammatory biomarkers, and left ventricular structure and function, COXPH = Cox Proportional Hazards Regression Model, RSF = Random Survival Forest, lSVM = Linear Support Vector Machines, nMTLR = Neural Multi-Task Logistic Regression, DeepSurv = Non-Linear Cox Proportional Hazards Deep Neural Network.

**Figure Legends:**

**Supplemental Figure 1:** Correlation matrix for all features used in the study. To avoid multicollinearity, features with a correlation coefficient greater than 0.80 were removed from further analyses, except for one feature in each set of correlations.

**Supplemental Figure 2:** Concordance index for feature combinations using traditional statistics. **(A)** Cox proportional hazards of feature combinations across time, evaluated through concordance index. Without splitting the data, MAE and mortality were assessed. MAE = major adverse events, including all-cause mortality and cardiovascular outcomes, Time in Days = time in days to the predicted event, PCE ASCVD = Pooled Cohort Equation estimated 10-year risk for atherosclerotic cardiovascular disease, Baseline Model = traditional risk factors, Biomarkers = inflammatory biomarkers, Imaging = left ventricular structure and function, All Features = traditional risk factors, inflammatory biomarkers, and left ventricular structure and function

**Supplemental Figure 3:** Parameters for machine and deep learning algorithms. **(A)** The fit methods for applying the external validation set to the trained algorithms of the internal training/testing sets. All fit methods are from the basic configurations of the PySurvival toolbox and were not optimized, with the exception of the RandomSurvivalForestModel, which was altered to build 500 trees. Hidden layers for the deep learning algorithms **(B)** nMTLR (no hyperparameter optimization) and **(C)** DeepSurv (hyperparameter optimization). COXPH = Cox Proportional Hazards Regression Model, RSF = Random Survival Forest, lSVM = Linear Support Vector Machines, nMTLR = Neural Multi-Task Logistic Regression, DeepSurv = Non-Linear Cox Proportional Hazards Deep Neural Network.

**Supplemental Figure 4:** Cumulative/dynamic time-dependent AUCs for the top five predictive features for MAE. Time-dependent AUCs were captured over 100 data points along the survival timeline (solid lines with dots), with the mean of each feature’s AUC displayed (dotted line) for **(A)** COXPH, **(B)**, RSF, **(C)** lSVM, **(D)** nMTLR, and **(E)** DeepSurv. Data are statistically significant with a *P* ≤ 0.05 when denoted by a “*****”. MAE = major adverse events, including all-cause mortality and cardiovascular outcomes, COXPH = Cox Proportional Hazards Regression Model, RSF = Random Survival Forest, lSVM = Linear Support Vector Machines, nMTLR = Neural Multi-Task Logistic Regression, DeepSurv = Non-Linear Cox Proportional Hazards Deep Neural Network.

**Supplemental Figure 5:** Cumulative/dynamic time-dependent AUCs for the top five predictive features for mortality. Time-dependent AUCs were captured over 100 data points along the survival timeline (solid lines with dots), with the mean of each feature’s AUC displayed (dotted line) for **(A)** COXPH, **(B)**, RSF, **(C)** lSVM, **(D)** nMTLR, and **(E)** DeepSurv. Data are statistically significant with a *P* ≤ 0.05 when denoted by a “*****”. COXPH = Cox Proportional Hazards Regression Model, RSF = Random Survival Forest, lSVM = Linear Support Vector Machines, nMTLR = Neural Multi-Task Logistic Regression, DeepSurv = Non-Linear Cox Proportional Hazards Deep Neural Network.

**Supplemental Figure 6:** Cumulative/dynamic time-dependent AUCs for the worst five predictive features for MAE. Time-dependent AUCs were captured over 100 points along the survival timeline (solid lines with dots), with the mean of each feature’s AUC displayed (dotted line) for **(A)** COXPH, **(B)**, RSF, **(C)** lSVM, **(D)** nMTLR, and **(E)** DeepSurv. Data are statistically significant with a *P* ≤ 0.05 when denoted by a “*****”. MAE = major adverse events, including all-cause mortality and cardiovascular outcomes, COXPH = Cox Proportional Hazards Regression Model, RSF = Random Survival Forest, lSVM = Linear Support Vector Machines, nMTLR = Neural Multi-Task Logistic Regression, DeepSurv = Non-Linear Cox Proportional Hazards Deep Neural Network.

**Supplemental Figure 7:** Cumulative/dynamic time-dependent AUCs for the worst five predictive features for mortality. Time-dependent AUCs were captured over 100 points along the survival timeline (solid lines with dots), with the mean of each feature’s AUC displayed (dotted line) for **(A)** COXPH, **(B)**, RSF, **(C)** lSVM, **(D)** nMTLR, and **(E)** DeepSurv. Data are statistically significant with a *P* ≤ 0.05 when denoted by a “*****”. COXPH = Cox Proportional Hazards Regression Model, RSF = Random Survival Forest, lSVM = Linear Support Vector Machines, nMTLR = Neural Multi-Task Logistic Regression, DeepSurv = Non-Linear Cox Proportional Hazards Deep Neural Network.
